# Preserved and reduced ejection fraction manifest as two mechanistically unique phenotypes of diastolic dysfunction

**DOI:** 10.1101/2025.08.17.25332552

**Authors:** Hayden McColl, Morgan Brookes, Michelle Price, Elias Fulthorp, Michele McGrady, Sean Lal, Martin Ugander

## Abstract

**Aims:** Diastolic dysfunction is commonplace in both heart failure with preserved (pEF) and reduced (rEF) ejection fraction (EF). However, differences in treatment response suggest that these classifications represent two unique phenotypes of diastolic dysfunction. We aimed to evaluate if patients with pEF and rEF would exhibit different mechanics of diastolic dysfunction.

**Methods and Results:** Patients undergoing echocardiography with normal EF and normal diastolic function, or grade 2 or 3 diastolic dysfunction with pEF or rEF were retrospectively identified. The mechanistic properties of diastolic function were assessed using the parameterised diastolic filling (PDF) method, which quantifies diastolic mechanics using the early mitral inflow pattern of the E-wave from Doppler echocardiography. Patients with diastolic dysfunction and pEF (n=75) or rEF (n=75) did not differ in E/e’, tricuspid regurgitation peak velocity, or left atrial volume index (p>0.05 for all). Compared to controls (n=75), left ventricular stiffness did not differ in either pEF or rEF (p>0.3 for all), while damping was increased in pEF (p<0.001), but reduced in rEF (p=0.005).

**Conclusions:** While pEF and rEF do not differ in key conventional echocardiographic measures of diastolic dysfunction, they are two unique physiologic conditions where the diastolic dysfunction manifests fundamentally differently. These findings challenge traditional assumptions about the role of myocardial stiffness in ventricular filling and underscore the importance of damping as a key mechanistic component of diastolic dysfunction. These insights may help explain differing responses to therapy across heart failure phenotypes and prompt further research into the targetable pathophysiological features that cause these changes.

## Introduction

Left ventricular diastolic dysfunction is a term used to describe abnormalities in ventricular relaxation. Abnormalities in diastolic function are present in a range of different cardiac conditions including both heart failure with reduced ejection fraction (HFrEF) and heart failure with preserved ejection fraction (HFpEF)^1^. Recent studies and guidelines suggest that diastolic dysfunction is not only near universal in both HFpEF and HFrEF but also has strong prognostic significance in these cohorts^2–4^. Diastolic dysfunction is commonly identified and graded using the 2025American Society of Echocardiography guidelines^3^. This approach combines measures of mitral annular tissue velocities, left atrial volumes, and the assessment of transmitral inflow Doppler profiles. Whilst useful for diagnosis, this grading system is not designed to assess the mechanisms which underlie diastolic dysfunction. As such, alternate methods of analysing ventricular filling have been proposed to better understand left ventricular (LV) diastolic function.

The parameterised diastolic filling (PDF) formalism is a method which accurately models early diastolic filling measured via pulsed wave (PW) Doppler echocardiography as a case of a damped harmonic oscillator^5^. A damped harmonic oscillator is a system analogous to a spring which experiences a proportionate restorative force when displaced from its resting state. When treating left ventricular filling as such, the velocity of the ‘spring’ over time can be quantified in terms of constants for ventricular stiffness, damping, and load. The PDF method involves fitting a curve to the mitral valve E-wave PW Doppler signal to solve for these three constants^6^. Key PDF parameters and their explanations are outlined in Table 1.

**Table 1.**
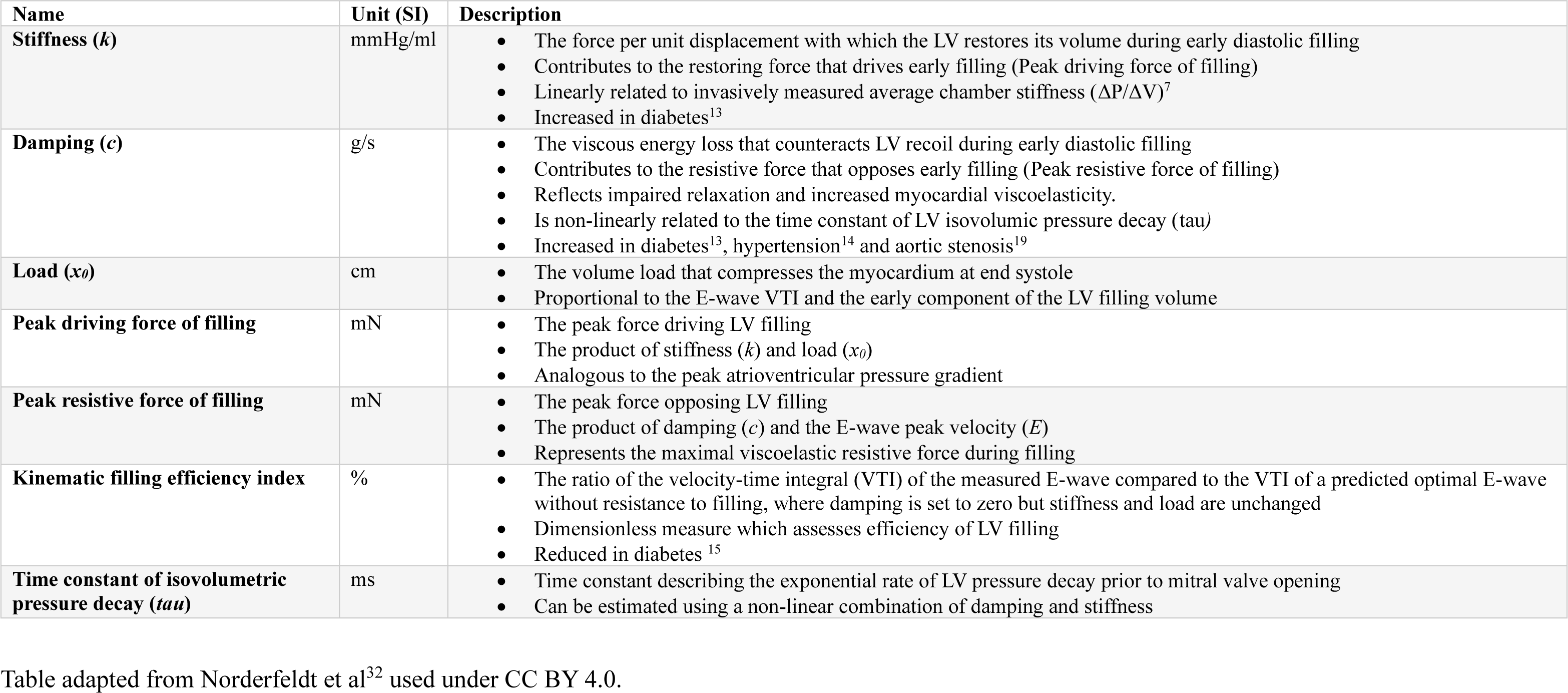
Description of key parameterized diastolic filling method parameters.

| Name | Unit (SI) | Description |
| --- | --- | --- |
| **Stiffness ($k$)** | mmHg/ml | The force per unit displacement with which the LV restores its volume during early diastolic filling<br>Contributes to the restoring force that drives early filling (Peak driving force of filling)<br>Linearly related to invasively measured average chamber stiffness ($\Delta P/\Delta V$)<sup>7</sup><br>Increased in diabetes<sup>13</sup> |
| **Damping ($c$)** | g/s | The viscous energy loss that counteracts LV recoil during early diastolic filling<br>Contributes to the resistive force that opposes early filling (Peak resistive force of filling)<br>Reflects impaired relaxation and increased myocardial viscoelasticity.<br>Is non-linearly related to the time constant of LV isovolumic pressure decay ($\tau$)<br>Increased in diabetes<sup>13</sup>, hypertension<sup>14</sup> and aortic stenosis<sup>19</sup> |
| **Load ($x_0$)** | cm | The volume load that compresses the myocardium at end systole<br>Proportional to the E-wave VTI and the early component of the LV filling volume |
| **Peak driving force of filling** | mN | The peak force driving LV filling<br>The product of stiffness ($k$) and load ($x_0$)<br>Analogous to the peak atrioventricular pressure gradient |
| **Peak resistive force of filling** | mN | The peak force opposing LV filling<br>The product of damping ($c$) and the E-wave peak velocity ($E$)<br>Represents the maximal viscoelastic resistive force during filling |
| **Kinematic filling efficiency index** | % | The ratio of the velocity-time integral (VTI) of the measured E-wave compared to the VTI of a predicted optimal E-wave without resistance to filling, where damping is set to zero but stiffness and load are unchanged<br>Dimensionless measure which assesses efficiency of LV filling<br>Reduced in diabetes<sup>15</sup> |
| **Time constant of isovolumetric pressure decay ($\tau$)** | ms | Time constant describing the exponential rate of LV pressure decay prior to mitral valve opening<br>Can be estimated using a non-linear combination of damping and stiffness |

Table adapted from Norderfeldt et al<sup>32</sup> used under CC BY 4.0.

In the PDF framework, stiffness refers to the ability of the LV to resist deformation and contributes to the force driving ventricular recoil in early ventricular filling. The stiffness constant has a linear correlation with invasively measured LV stiffness (ΔP/ΔV)^7^. In contrast, damping reflects viscoelastic energy loss and contributes to the force which opposes ventricular filling. This is analogous to a damper or dashpot, a mechanical device that is commonly used to dampen the closing velocity of a door that closes by a spring mechanism. Finally, load represents the deforming volume on the ventricle which occurs during contraction in systole^5^, and load is proportional to the velocity-time integral of the E-wave.

Diastolic dysfunction is a key component in the pathophysiology and development of symptoms in both HFpEF and HFrEF and has prognostic importance in both these conditions^1, 4^. Whilst diastolic dysfunction is ubiquitous in both these conditions, treatment options and treatment efficacy differ greatly and an understanding of the mechanisms driving these differences is currently lacking. The aim of this study was to investigate mechanistic differences in patients with diastolic dysfunction and preserved ejection fraction (pEF) or reduced ejection fraction (rEF) using the PDF method. We hypothesized that diastolic dysfunction in these conditions represents two mechanistically unique disease phenotypes.

## Methods

### Study Design

This single-centre, retrospective study compared control patients with normal systolic and diastolic function to patients with diastolic dysfunction and preserved or reduced ejection fraction. All included patients were referred for cardiac assessment at a large academic out-patient cardiology practice between January 2018 and December 2023. The study was performed according to the Declaration of Helsinki and ethics approval was received from the Sydney Local Health District Ethics Board (X19-0286 & 2019/ETH12064) as well as the human research ethics committee of the participating centre, with a retrospective waiver of informed consent.

Patients aged >50 years who underwent transthoracic echocardiography were eligible for inclusion in this study. Patients were excluded if they had moderate or greater valvular disease, hypertrophic cardiomyopathy or cardiac surgery including valvular replacement, coronary artery bypass grafting or interventions that would disrupt the pericardium. Patients were additionally excluded in the cases where insufficient transmitral Doppler signals were able to be obtained. Figure 1 outlines the patient inclusion flowchart.

**Figure 1.**
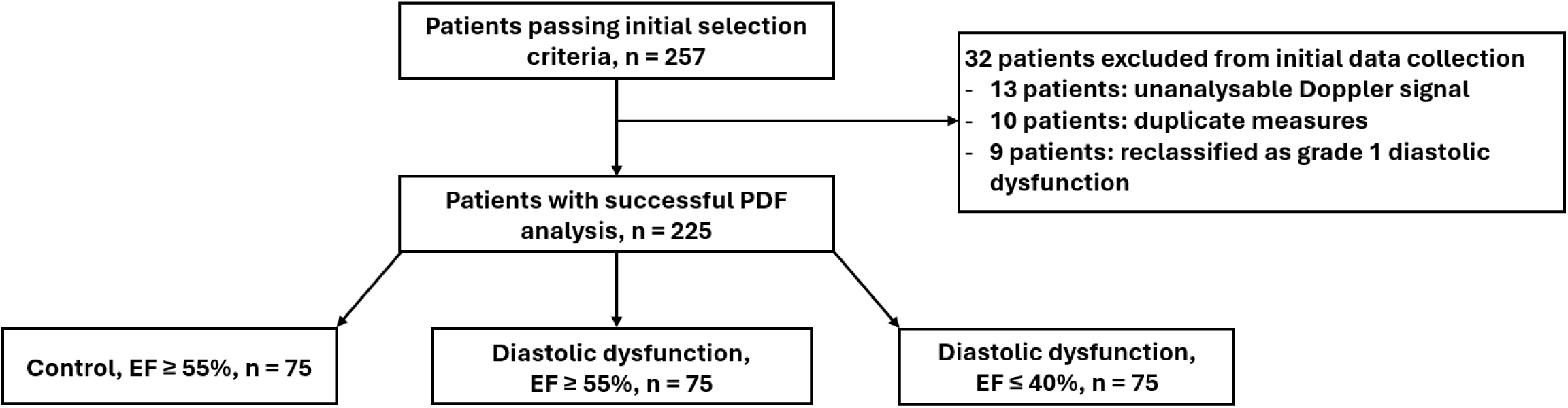
Patient inclusion flowchart.

Echocardiographic analysis was performed using a commercially available clinical echocardiography system (VIVID, GE Healthcare, Chicago, Illinois, USA) in accordance with the American Society of Echocardiography (ASE) guidelines for a comprehensive transthoracic echocardiographic examination^8^. Patients were classified as having either normal or impaired diastolic function. Impaired diastolic function was defined as the presence of either grade 2 or 3 diastolic dysfunction as per the 2016 ASE guidelines^2^.

### Parameterised Diastolic Filling Analysis

PDF analysis was performed using the web-based application, Echo E-waves (www.echoewaves.org)^6^. A curve-fit of the velocity profile of the PW doppler signal of early diastolic fitting was estimated using measures of peak E-wave velocity, E-wave acceleration time, and E-wave deceleration time according to validated methods^9^. From this, the PDF parameters for stiffness, damping and load were derived, in addition to secondary calculated parameters for the estimated time constant of isovolumic relaxation (tau), kinematic filling efficiency index (KFEI), filling energy, peak driving force, peak resistive force and damping index^6^. Figure 2 outlines the workflow for PDF analysis.

**Figure 2.**
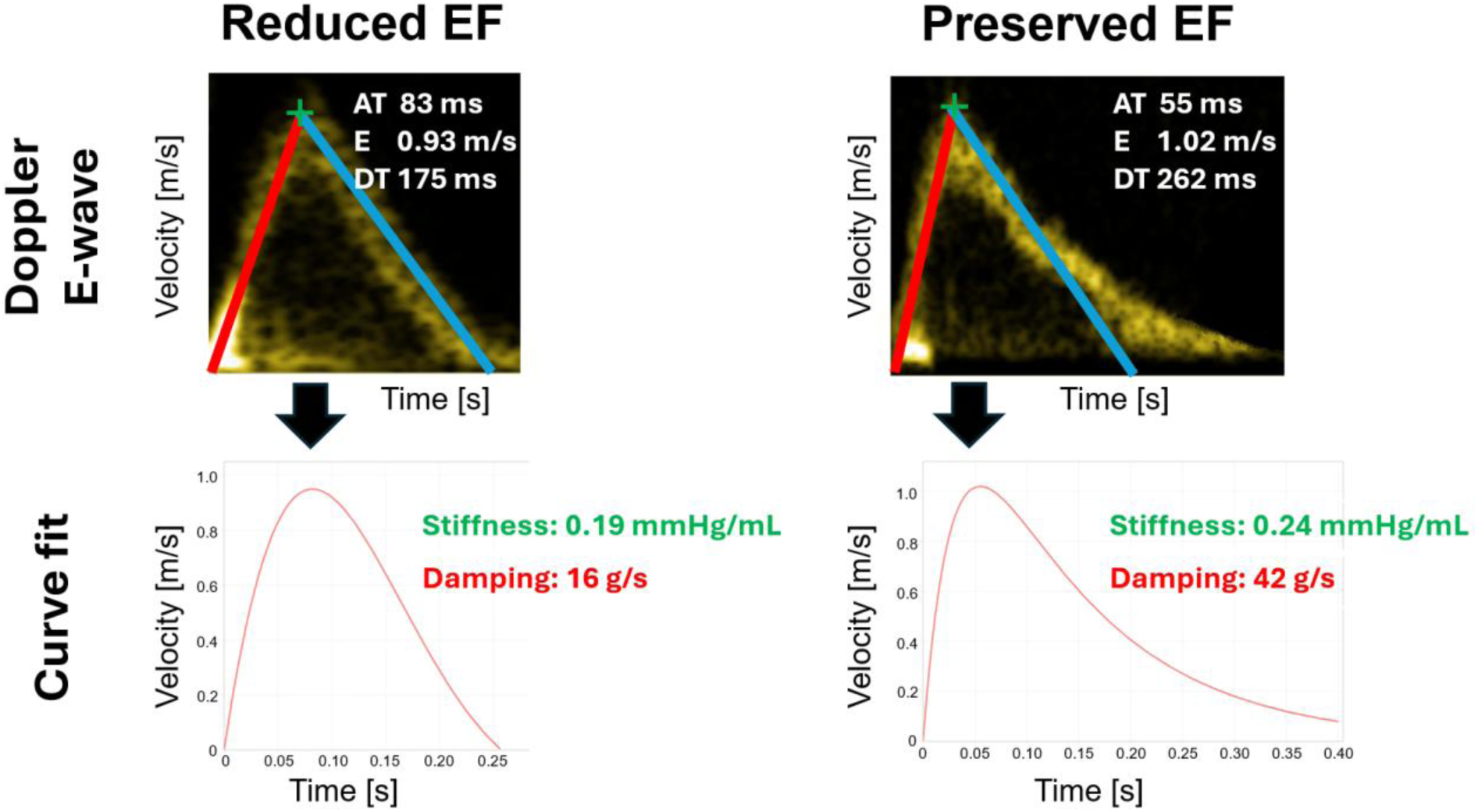
Parameterized diastolic filling methodology. Representative cases of patients with diastolic dysfunction and reduced (left) or preserved (right) ejection fraction (EF). Top row shows pulsed-wave Doppler recording of early transmitral blood flow velocities (E-wave) over time. The red line denotes the acceleration time (AT), the green marker denotes the peak velocity (E), and the blue line denotes the deceleration time (DT). These three measures together uniquely describe the E-wave (Bottom row) that can be defined mathematically according to the expected behaviour of left ventricular filling in terms of a damped harmonic oscillator.

### Statistical Analysis

Statistical analysis was conducted using IBM SPSS Statistics, version 29. Continuous variables were compared using either one-way ANOVA or Kruskal-Wallis methods depending on the distribution. Continuous variables are reported as mean ± standard deviation or median [interquartile range] as appropriate.

## Results

The study population consisted of 225 patients; 75 control patients with normal systolic and diastolic function, and 150 with abnormal diastolic function (grade 2 or 3 diastolic function). Patients with impaired diastolic function were further subdivided into either preserved (EF>55%, n=75) or reduced (EF<40%, n=75) left ventricular ejection fraction groups. The baseline characteristics of the 3 groups are presented in Table 2. Patients with diastolic dysfunction and pEF or rEF did not differ in the ratio of early transmitral blood flow to early myocardial velocity (E/e’), left atrial volume index (LAVI) or tricuspid regurgitation maximum blood flow velocity (TR Vmax). E peak velocity and average e’ was increased in patients with diastolic dysfunction and pEF compared with rEF (87 [72-97] vs 77 [65-91] cm/s, p=0.009, and 5.8 [4.8-7.0] vs 4.1 [3.5-5.7] cm/s, p=0.001, respectively). The early-to-atrial transmitral blood flow velocity ratio (E/A) was decreased in patients with diastolic dysfunction and pEF compared to rEF (0.87 [0.70-1.14] vs 1.60 [1.00-2.27], p<0.001).

**Table 2.**
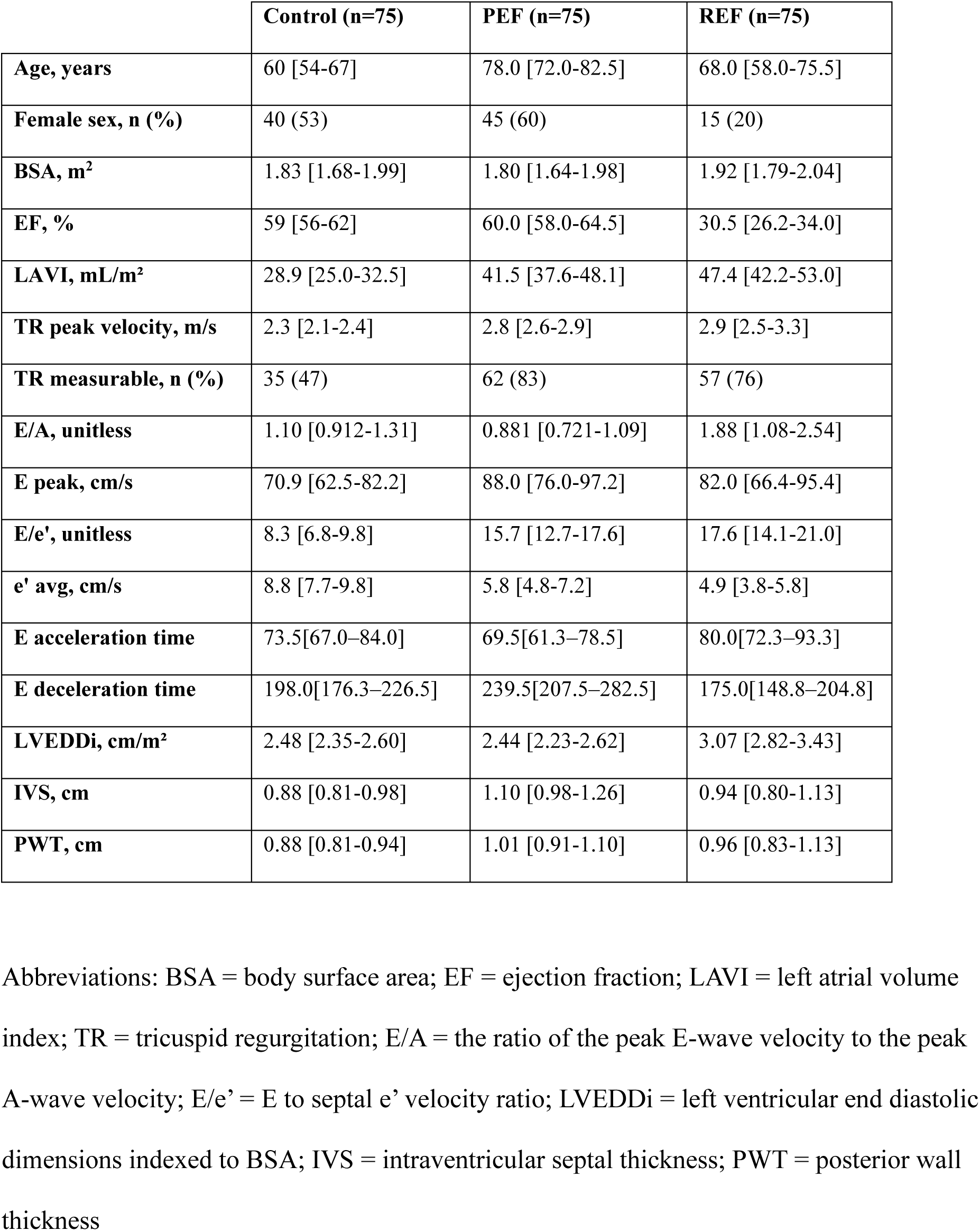
Patient characteristics of the control, preserved and reduced ejection fraction cohorts.

|  | **Control (n=75)** | **PEF (n=75)** | **REF (n=75)** |
| --- | --- | --- | --- |
| **Age, years** | 60 [54-67] | 78.0 [72.0-82.5] | 68.0 [58.0-75.5] |
| **Female sex, n (%)** | 40 (53) | 45 (60) | 15 (20) |
| **BSA, m<sup>2</sup>** | 1.83 [1.68-1.99] | 1.80 [1.64-1.98] | 1.92 [1.79-2.04] |
| **EF, %** | 59 [56-62] | 60.0 [58.0-64.5] | 30.5 [26.2-34.0] |
| **LAVI, mL/m<sup>2</sup>** | 28.9 [25.0-32.5] | 41.5 [37.6-48.1] | 47.4 [42.2-53.0] |
| **TR peak velocity, m/s** | 2.3 [2.1-2.4] | 2.8 [2.6-2.9] | 2.9 [2.5-3.3] |
| **TR measurable, n (%)** | 35 (47) | 62 (83) | 57 (76) |
| **E/A, unitless** | 1.10 [0.912-1.31] | 0.881 [0.721-1.09] | 1.88 [1.08-2.54] |
| **E peak, cm/s** | 70.9 [62.5-82.2] | 88.0 [76.0-97.2] | 82.0 [66.4-95.4] |
| **E/e', unitless** | 8.3 [6.8-9.8] | 15.7 [12.7-17.6] | 17.6 [14.1-21.0] |
| **e' avg, cm/s** | 8.8 [7.7-9.8] | 5.8 [4.8-7.2] | 4.9 [3.8-5.8] |
| **E acceleration time** | 73.5[67.0–84.0] | 69.5[61.3–78.5] | 80.0[72.3–93.3] |
| **E deceleration time** | 198.0[176.3–226.5] | 239.5[207.5–282.5] | 175.0[148.8–204.8] |
| **LVEDDi, cm/m<sup>2</sup>** | 2.48 [2.35-2.60] | 2.44 [2.23-2.62] | 3.07 [2.82-3.43] |
| **IVS, cm** | 0.88 [0.81-0.98] | 1.10 [0.98-1.26] | 0.94 [0.80-1.13] |
| **PWT, cm** | 0.88 [0.81-0.94] | 1.01 [0.91-1.10] | 0.96 [0.83-1.13] |

Abbreviations: BSA = body surface area; EF = ejection fraction; LAVI = left atrial volume index; TR = tricuspid regurgitation; E/A = the ratio of the peak E-wave velocity to the peak A-wave velocity; E/e' = E to septal e' velocity ratio; LVEDDi = left ventricular end diastolic dimensions indexed to BSA; IVS = intraventricular septal thickness; PWT = posterior wall thickness

Results are summarized graphically in figure 3. Stiffness did not differ between controls (0.13 [0.06-0.20] mmHg/ml) and pEF (0.12 [0.06-0.18] mmHg/ml, p=1.0) or rEF (0.11 [0.05-0.17] mmHg/ml, p=0.33). Compared to controls, damping was increased in pEF (24.1 [20.5-33.2] vs 19.8 [15.1-24.8] g/s, p<0.001) and reduced in rEF (16.1 [12.5-19.8] g/s, p=0.005). Compared to controls, load was increased in pEF (13.7 [11.3-16.8] vs 9.7[8.5-11.2]cm, p<0.001) but did not differ in patients with rEF (10.4 [8.9-12.0]cm, p=0.22).

**Figure 3.**
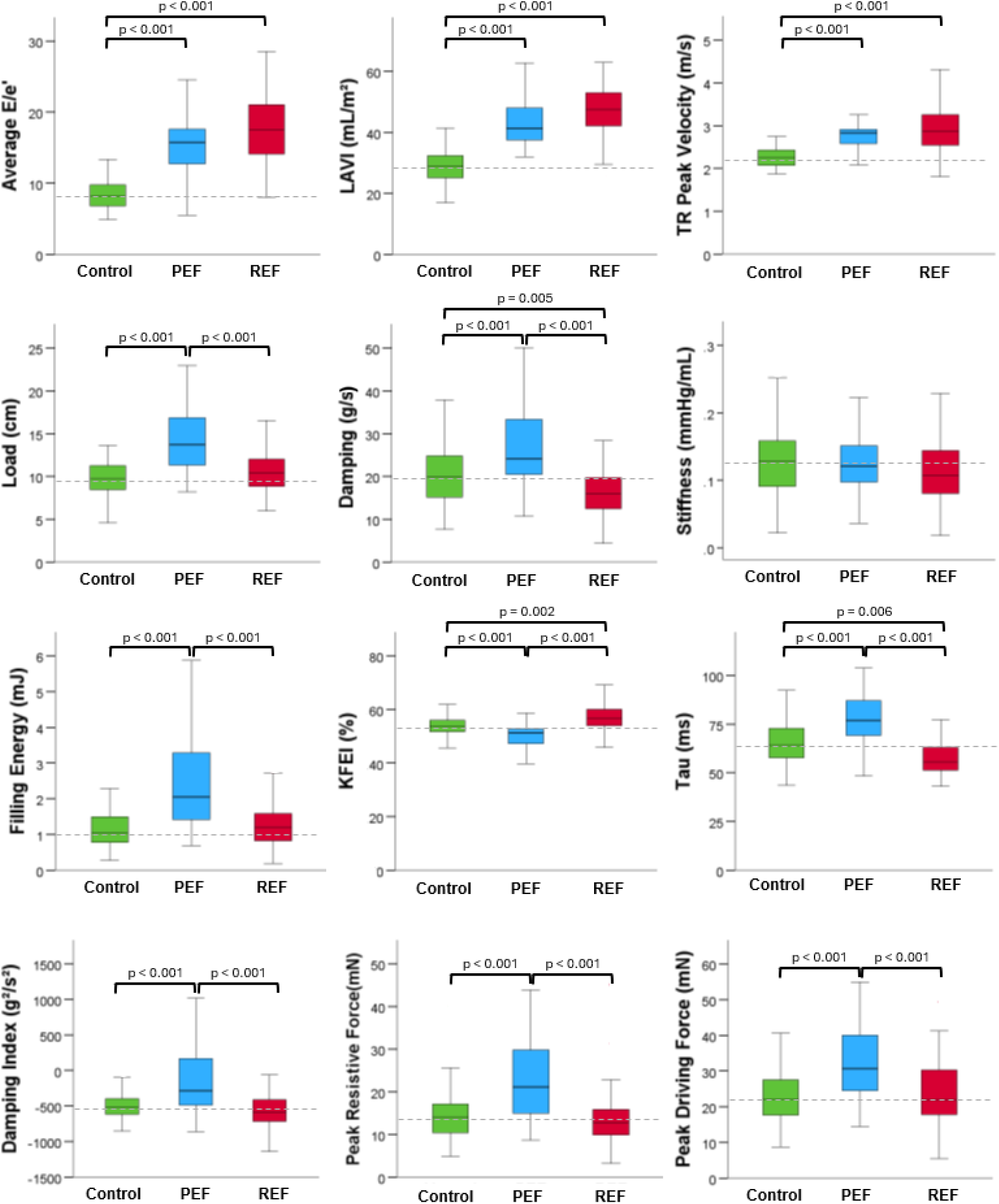
Conventional echocardiographic and parameterized diastolic filling parameters of controls, preserved, and reduced ejection fraction groups. Brackets denote statistical significance where present. Horizontal grey dotted line indicates median value for control cohort. Abbreviations: TR = tricuspid regurgitation. KFEI = kinematic filling efficiency index

Compared to controls, estimated tau was increased in patients with pEF (76 [69-87] vs 64 [58-73] ms, p<0.001) and reduced in rEF (56 [51-63] ms, p=0.006). Conversely, KFEI was decreased in pEF (51.2 [47.3-52.7] vs 53.7 [51.7-56.0] %, p<0.001) but increased in rEF (56.7 [54.1-60.1] %, p=0.002). Comprehensive numerical results are detailed in Appendix Table 1.

## Discussion

The main finding of this study is that despite similar diastolic dysfunction grading using conventional echocardiographic guidelines, diastolic dysfunction in patients with pEF represents a fundamentally different mechanistic phenotype to that of patients with rEF.

The mechanisms which govern LV filling are complex and represent an interplay between intrinsic properties of the myocardium, pericardial physiology, and hemodynamic loading conditions^10^. By modelling this process as a case of damped harmonic motion we gain an appreciation of the mechanistic differences between phenotypes of diastolic dysfunction. A clear appreciation of the definitions and application of key parameters, particularly the primary parameters of the PDF method (stiffness, damping and load) is essential for meaningful interpretation of these results.

Diastolic dysfunction and HFpEF is commonly described as being due to a “stiff” ventricle. However, the term stiffness is often poorly defined and is inconsistently used throughout literature. In mechanistic terms, stiffness is a measure of an object’s ability to resist deformation in response to an applied force^11^. The stiffer an object, like the myocardium of the left ventricle, the more force which is required to deform it. Consequently, this also means that a stiffer ventricle will exert a greater force to recoil to its resting shape. When we apply this to early diastole, the stiffness of myocardium *aids* ventricular relaxation and filling as the compressed myocardium at end-systole returns to a relative resting state during diastasis.

By comparison, damping represents viscoelastic energy loss. Viscoelasticity is a common property of biological tissues, including myocardium, which describes the presence of both elastic elements which allow the ventricle to deform and recoil, as well as viscous elements which generates velocity-dependent resistance to motion. When modelling diastole, viscoelastic energy loss represents energy dissipated, commonly as heat, during ventricular relaxation^12^. In contrast to stiffness, the damping properties of the myocardium contribute to the force which *opposes* ventricular filling.

Load represents the volume of displacement from the resting state. It does not have a direct comparison in the myocardium, however, is linearly related to the E-wave velocity-time integral and likely reflects volumetric preload^5, 6^. Taken together, applying these definitions aids in understanding the findings of this study.

Differences in damping emerged as a key driver of the distinct diastolic dysfunction phenotypes observed in patients with preserved versus reduced ejection fraction. Patients with diastolic dysfunction and pEF had higher values of damping when compared with controls. Increased damping has also been noted in hypertensive heart disease and patients with diabetes, both thought to be a potential precursors for overt diastolic dysfunction^13,14^. In contrast, we found that damping is *decreased* in HFrEF compared to controls. Damping is velocity-dependent, and potentially reduced systolic contraction in rEF patients generates less stored elastic energy and subsequent velocity, resulting in overall less impact of damping during ventricular filling. The kinematic filling efficiency index (KFEI) is a measure derived from PDF analysis that similarly highlights the impact of damping on diastolic function. This dimensionless measure is the ratio of the measured PDF contour-fit E-wave, to a predicted optimal E-wave which keeps the same load and stiffness but sets damping to zero^15^. Patients with pEF have worse KFEI than controls, suggesting a greater degree of diastolic impairment. This is in line with prior research demonstrating impaired KFEI in patients with diabetes, a condition which has been linked with the development of HFpEF^15^.

One key finding of our study is that stiffness did not differ between the control, pEF, and rEF groups. This is an observation that contrasts prior studies which suggest that patients with diastolic dysfunction may have increased chamber stiffness ^16, 17^. This finding is most likely due to the different methods by which myocardial stiffness has been assessed. These prior studies commonly quantify stiffness through instantaneous measures of the pressure-volume relationship at end diastole. Diastolic filling is a dynamic process and may not be optimally assessed through measures at only a single time point, even using methodology which averages these measures and/or incorporates assessment during different loading conditions. It is worth noting that stiffness as assessed by the PDF method has been shown to correlate linearly with invasive measures of average chamber stiffness measured as the change in LV pressure with respect to a change in LV volume (ΔP/ΔV)^7^. Moreover, the PDF method quantifies diastolic mechanics over the entirety of early ventricular filling. Also, inherent to the PDF method is the quantification of both stiffness and damping. As shown in the current results, damping is a physical property of the myocardium that is changed in diastolic dysfunction, and which has largely been overlooked in the literature owing to lack of methods for quantification^5^. Taken together, the current manuscript may offer a more nuanced assessment of diastolic function compared with purely instantaneous measures.

Load, and its closely related parameter peak driving force, was elevated in those with pEF but not with rEF when compared to controls. Load has a close linear relationship with the E-wave velocity-time integral and is representative of the volumetric contribution of early diastolic filling^7^. Peak driving force represents the peak transmitral pressure gradient and correlates with invasive haemodynamic measurements^6^. Increased load in pEF may reasonably reflect increased relative contribution of early diastolic filling, which is a known phenomenon with progressive diastolic dysfunction represented by increase in the E/A ratio^18^. Increased load and damping has been noted in aortic stenosis populations, which is another heart failure phenotype which can manifest as HFpEF with ventricular remodelling driven by increased afterload^19^. Notably, damping is not limited to relaxation but also contributes velocity-dependent resistance during both systole and diastole. The increased load observed in pEF may reflect the greater energy required during systole to overcome the heightened damping properties of the myocardium in pEF. Although the precise mechanisms remain unclear, these findings suggest that patients with pEF may experience elevated left ventricular filling pressures, potentially exceeding those seen in rEF.

This study demonstrates that diastolic dysfunction with preserved or reduced ejection fraction are unique mechanistic entities that in many ways manifest with deviations from controls that go in opposite directions. The reasons for this are likely complex and multifactorial. This may be related to differences in ventricular remodelling at a cellular level resulting in global structural changes. In HFpEF, cardiomyocytes have been shown to exhibit transverse hypertrophy whilst keeping a relatively fixed cell length. This subsequently results in the concentric remodelling seen in numerous HFpEF phenotypes^20^. In comparison, HFrEF is commonly due to cardiomyocyte loss with resultant chamber dilation and eccentric remodelling. At a cellular level, there is relative sparsity of cardiomyocytes with elongation in longitudinal dimensions resulting on overall chamber dilation^21^. Myocardial fibrosis is increased in both HFpEF and HFrEF, though the underlying patterns differ^22^. HFpEF is typically characterised by diffuse myocardial fibrosis with expansion of the extracellular matrix, often occurring in the absence of significant cardiomyocyte loss^23^. In contrast, HFrEF more commonly exhibits dense replacement myocardial fibrosis resulting from scarring due to focal cardiomyocyte injury and loss with the typical example being chronic myocardial infarction^22^. These distinct patterns of diffuse as opposed to focal myocardial fibrosis likely impact diastolic function differently and may contribute to the observed differences between the two phenotypes. Finally, molecular differences including modification in titin protein isoform and intracellular calcium sequestration have also been noted between HFpEF and HFrEF^24^. Conceptually, increased damping could be related to diffuse myocardial fibrosis, disruption of the normal myofiber organisation, impaired myocardial calcium sequestration, or abnormalities in sarcomere cross-bridge uncoupling^15, 25, 26^. In contrast, stiffness is generated by a multitude of intrinsic myocardial factors. Whilst the abnormal accumulation of collagen seen in diffuse myocardial fibrosis contributes to damping, the innate elastic properties of the normal extracellular matrix, primarily driven by elastin help generate myocardial stiffness^27^. Intracellular titin and cytoskeletal microtubules have also been shown to modulate myocardial stiffness and contribute to ventricular filling^28, 29^. Finally, stiffness is also driven by external factors such as pericardial characteristics and the influence of the great vessels on early ventricular recoil^29, 30^.

There are several limitations to the present study. First, the PDF methodology is sensitive to the precise measurement of the components of early diastolic mitral inflow (E-wave), specifically the acceleration time and deceleration time. Small variations in measured acceleration time and deceleration time can result in relatively large differences in derived PDF parameters, thus introducing measurement variability. This sensitivity may limit the clinical utility of the PDF for discriminating between different manifestations of diastolic function at the individual patient level. However, there are available published normal reference values^31^. Notably, this measurement variability does not limit the ability to glean mechanistic insight from group comparisons in sufficiently powered sample sizes such as in the current study. Secondly, while the PDF approach offers mechanistic insight, diastolic function is inherently complex and influenced by multiple dynamic factors. As with traditional echocardiographic parameters, PDF measures are susceptible to changes in preload and afterload, which may confound interpretation. The inherent variability in loading conditions does limit the current utility of PDF analysis on an individual patient diagnostic level. However, we are still able to gain a deeper understanding of overall mechanics when looking at cohorts of patients. Future studies using the PDF methodology could consider paired TTE and brain natriuretic peptide measurements to further correct for this variation.

Thirdly, both HFpEF and HFrEF are two broad classifications of heart failure phenotypes that encompass a heterogenous population of different contributory pathologies. The differences observed between the pEF and rEF groups in the current study may be driven by a dominant disease phenotype within the cohort and may not be generalisable to all aetiologies of heart failure. As data on comorbidities were not available, we are unable to explore these differences. Despite these shortcomings, the current study represents the first application of any method that can quantitatively show the mechanistic differences in diastolic dysfunction between pEF and rEF, thus opening for future prospective studies to better understand the clinical applications of these differences. Finally, the PDF methodology as employed in this study is confined to analysis of the E-wave and therefore does not account for the atrial contribution to diastolic filling. Abnormalities manifesting during late diastole, including those related to atrial dysfunction or increased ventricular stiffness during atrial contraction, are not captured.

## Conclusions

Although conventional echocardiographic grading may similarly classify diastolic dysfunction in patients with preserved and reduced ejection fraction, these groups exhibit distinct underlying mechanistic phenotypes. Importantly, the phenomenon of diastolic dysfunction appears to be driven by differences in myocardial damping, rather than stiffness (Graphical Abstract). A deeper understanding of the impact of structural myocardial alterations driving these changes may facilitate the development of targeted therapies, particularly for heart failure with preserved ejection fraction, where effective treatment options remain limited.

## Data Availability

The data underlying this article can be made available upon reasonable request to the corresponding author.

