## Supplementary material for "Preserved and reduced ejection fraction manifest as two mechanistically unique phenotypes of diastolic dysfunction"

**Supplementary information**

**Appendix Table 1. Comparison of parameterized diastolic filling parameters between control, preserved and reduced ejection fraction groups**

|  | **Normal (n=75)** | **PEF (n=75), p vs controls** | **REF (n=75), p vs control** | **p, PEF vs REF** |
| --- | --- | --- | --- | --- |
| **Stiffness, mmHg/mL** | 0.13 [0.06-0.20] | 0.12 [0.06-0.18], p=1.0 | 0.11 [0.05-0.17], p=0.33 | p=0.59 |
| **Damping, g/s** | 19.8 [15.1-24.8] | 24.1 [20.5-33.2], **p<0.001** | 16.1 [12.5-19.8], **p=0.005** | **p <0.001** |
| **Load, cm** | 9.7[8.5-11.2] | 13.7 [11.3-16.8], **p<0.001** | 10.4 [8.9-12.0], p=0.22 | **p<0.001** |
| **Tau, ms** | 64.0 [57.6-72.8] | 76.9 [69.2-87.1], **p<0.001** | 55.6 [51.4-63.1], **p=0.006** | **p<0.001** |
| **KFEI, %** | 53.7 [51.7-56.0] | 51.2 [47.3-52.7], **p<0.001** | 56.7 [54.1-60.1], **p=0.002** | **p<0.001** |
| **Peak driving force, mN** | 22.1 [17.6-27.6] | 30.7 [24.5-40.0], **p<0.001** | 22.0 [17.8-30.3], p=1.0 | **p<0.001** |
| **Peak resistive force, mN** | 14.1 [10.4-17.1] | 21.1 [15.0-29.8], **p<0.001** | 12.8 [9.93-15.9], p=1.0 | **p<0.001** |
| **Damping index, g²s²** | -509 [-618- -399] | -283 [-491-164], **p<0.001** | -583 [-714- -409], p=0.10 | **p<0.001** |
| **Filling energy, mJ** | 1.05 [0.78-1.49] | 2.04 [1.42-3.29], **p<0.001** | 1.21 [0.82-1.59], p=0.10 | **p<0.001** |
